# Rapid identification of Sars-CoV-2 variants of concern using the portable *peak*PCR platform

**DOI:** 10.1101/2021.05.21.21256124

**Authors:** Salome Hosch, Philippe Bechtold, Philipp Wagner, Amalia Ruiz-Serrano, Michele Gregorini, Denise Siegrist, Olivier Engler, Wendelin J. Stark, Claudia A. Daubenberger, Tobias Schindler

## Abstract

The need for tools which allow rapid detection and continuous monitoring of Sars-CoV-2 variants of concern (VOC) is greater than ever, as these variants are more transmissible and therefore increase the pressure of COVID-19 on healthcare systems. To address this demand, we aimed to develop and evaluate a robust and fast diagnostic approach for identification of Sars-CoV-2 VOC-associated spike gene mutations. Our diagnostic assays detect the E484K and N501Y SNPs as well as a spike gene deletion (HV69/70) and can be run on standard laboratory equipment or on the portable rapid diagnostic technology platform *peak*PCR. The assays achieved excellent diagnostic performance when tested with RNA extracted from culture-derived Sars-CoV-2 VOC lineages. Simplicity of usage and the relatively low costs are advantages which make our approach well-suited for decentralized and rapid testing, especially in resource limited settings.

## Introduction

More than a year after the World Health Organization (WHO) declared the severe acute respiratory syndrome coronavirus type 2 (Sars-CoV-2) outbreak a Public Health Emergency of International Concern, Coronavirus disease 2019 (COVID-19) has caused more than 3.4 million deaths ^1^. Public health systems globally are severely impacted and are further challenged by the emergence of Sars-CoV-2 variants carrying mutations that are of concern (VOC) ^2^. Molecular diagnostic tools, particularly reverse transcription quantitative polymerase chain reaction (RT-qPCR) for viral RNA detection and next-generation sequencing (NGS) for molecular monitoring Sars-CoV-2 genetic diversity at the whole genome level have proven critical for public health decision-making ^3^. Investigating Sars-CoV-2 genomes by NGS to track transmission chains, understand transmission dynamics and rapidly identify mutations that potentially have an impact on transmissibility, morbidity and mortality as well as escape diagnostics or vaccine induced immunity have become an integral part of public health measures during this pandemic ^4^.

Since the first whole genome sequence (WGS) analysis of Sars-CoV-2 has been published in January 2020 ^5^, the virus has been continuously sequenced, characterized and made publicly available through global initiatives such as Global Initiative on Sharing All Influenza Data (GISAID). More than 1.6 million Sars-CoV-2 sequences have been publicly shared via GISAID and numerous mutations in the gene encoding the spike protein have been identified ^6^. For example, the D614G variant has been shown to increase the viral load of infected patients and has replaced the original variant since June 2020 around the globe ^7^. More recently, Sars-CoV-2 lineages characterized by a combination of multiple mutations in the spike gene have emerged independently in different regions of the world. The Sars-CoV-2 lineages B.1.1.7 (also known as VOC 202012/01 or 501Y.V1), B.1.351 (also known as 501Y.V2) and P.1 (also known as B.1.1.28.1 or 501Y.V3) are currently attracting most attention ^8^. B.1.1.7 was first described in mid-December 2020 in the United Kingdom, the mutation appears to have substantially increased transmissibility and has quickly developed into the dominant variant circulating in the UK and beyond ^9^. B.1.351 was identified in December 2020, emerged most likely in South Africa and is also associated with higher transmissibility ^10^. Lineage P.1 was identified in January 2021 in Manaus, the largest city in the Amazon region of Brazil ^11^. In January 2021, this region experienced an resurgence of COVID-19, despite the reported high seroprevalence of antibodies against Sars-CoV-2 in this population ^12^. It has been estimated that 76% of the population had been infected with SARS-CoV-2 by October 2020 ^13^.

To rapidly detect and continuously monitor the appearance, introduction and spread of (novel) VOCs, the level of molecular surveillance needs to be increased globally. The gold standard of genomic surveillance, NGS, allows unbiased identification of mutations, but is limited by its relatively slow sample-to-result turnaround time and level of laboratory infrastructure and expertise required. Furthermore, relatively high costs of NGS increase the financial burden on establishing a widespread VOC tracking strategy, a limiting factor particularly for resource-limited settings. There is a need for mutation-specific, cost-efficient diagnostic assays allowing high-throughput screening of a significant proportion of Sars-CoV-2 positive individuals to identify transmission dynamics of VOCs. Therefore, we have designed rapid and cost-effective RT-qPCR assays detecting relevant mutations in the spike protein of Sars-CoV-2. The N501Y mutation is found in all three major VOCs, while the E484K mutation is restricted to the B.1.351 and P.1 lineages. The B.1.1.7 lineage is characterized by an additional spike gene deletion (ΔHV69/70). To further simplify, decentralize and speed up the process of VOC identification, we transformed our assay to a portable and inexpensive qPCR device, named *peakPCR* ^14^. The device can complete up to 20 RT-qPCR reactions in less than 40 minutes. Important characteristic of *peak*PCR are the relatively low cost of production and the simplicity of usage. By using chips that are preloaded with lyophilized RT-qPCR reagents, the user interaction is reduced to loading the sample onto the chip. Furthermore, the preloaded *peak*PCR chips can easily be shipped and stored at room temperature, making cold chains redundant.

Here, we report the development of a new approach for rapid, robust and decentralized identification of Sars-CoV-2 variants of concern which can be run on standard laboratory RT-qPCR equipment as well as on the portable and rapid diagnostic technology platform *peak*PCR.

## Material and Methods

### Sars-CoV-2 cell culture supernatants and clinical samples for assay evaluation

The cultivation of SARS-CoV-2 was carried out in a Biosafety level 3 laboratory and conducted under appropriate safety conditions. Three different VOC lineages of Sars-CoV-2, namely B.1.1.7, B.1.351 and P.1 provided from University Hospital of Geneva, Laboratory of Virology were grown on VeroE6/TMPRSS2 cells obtained from the Centre For AIDS Reagents (National Institute for Biological Standards and Control) ^15,16^. The day before infection VeroE6/TMPRSS2 cells were seeded at 2×10^6^ cells per T75 flask in DMEM (Seraglob, Switzerland) supplemented with 10% FBS (Merck, Germany) and 2% SEeticin (Seraglob, Switzerland). At the day of infection, the cells reached about 70-90% confluency. The growth medium was removed and replaced with 5 mL of infection medium (DMEM+2% FBS+2% SEeticin). Cells were inoculated with 70 µL of SARS-CoV-2 swab material and incubated for 1 h at 37°C, 5% CO2, and >85% humidity. After the adsorption, 10 mL of infection medium was added to each flask. Cells were observed for cytopathic effects (CPE) for 3-6 days using EVOSTM FL Digital Inverted Microscope. When CPE reached 40-100% the supernatant was collected, cleared from cell debris by centrifugation (10 min at 500g) and samples were aliquoted and frozen. TCID50 was determined on VeroE6/TMPRSS2 cells. Virus was inactivated with Qiazol and RNA was extracted with RNeasy Plus Universal Mini Kit (Qiagen, Germany).

As a positive control and for initial assay evaluation a 1869 bp long synthetic Sars-CoV-2 spike gene fragment (genome position 21,557-23,434 bp), based on the sequence of B.1.1.7 was synthesized (Sequence is provided in *Appendix data 1)*. Using a serial dilution of the synthetic spike gene a calibration curve ranging from 0.05 to 50,000,000 copies/µL was prepared (data provided in *Appendix figure 1)*. The initial viral copy number per µL (cp/µL) of the cell culture derived RNA from Sars-CoV-2 was estimated using the calibration curve’s y-intercept and its slope. Serial dilutions of the RNA extracted from culture supernatants of Sars-CoV-2 isolates Wuhan Hu-1, B.1.1.7, B.1.351 and P.1, ranging from 0.1 to 1,000,000 cp/µL was prepared and used to evaluate the assays’ performance on both RT-qPCR platforms. Additionally, a Sars-CoV-2 RT-qPCR diagnostic assay, targeting the envelope (E) gene, published by the Institute of Virology at Charité (Berlin, Germany), was used to monitor the RNA integrity on both platforms^17^.

### Designing HV69/70-deletion-, E484K-and N501Y-specific RT-qPCR assays

We developed assays targeting the HV69/70-deletion, the E484K-, and the N501Y-single nucleotide polymorphisms (SNP). For standard RT-qPCR platforms like the Bio-Rad CFX96 device, multiplex assays were developed. The multiplex assays are able to detect both sequence variations, the wildtype and mutated, in a single RT-qPCR reaction. For rapid identification of mutations of interest for mobile and rapid RT-qPCR platforms, such as the *peak*PCR device, only the mutated sequence variation is detected and no multiplex amplification is performed. SNP discrimination was enhanced by using primers and probe containing Locked Nucleic Acids (LNAs). Sequence analysis and primer design were done using the Geneious Prime® 2021.0.3 software. All oligos, including the LNAs, were synthesized at Microsynth AG (Balgach, Switzerland) and provided in Table 1.

**Table 1.**
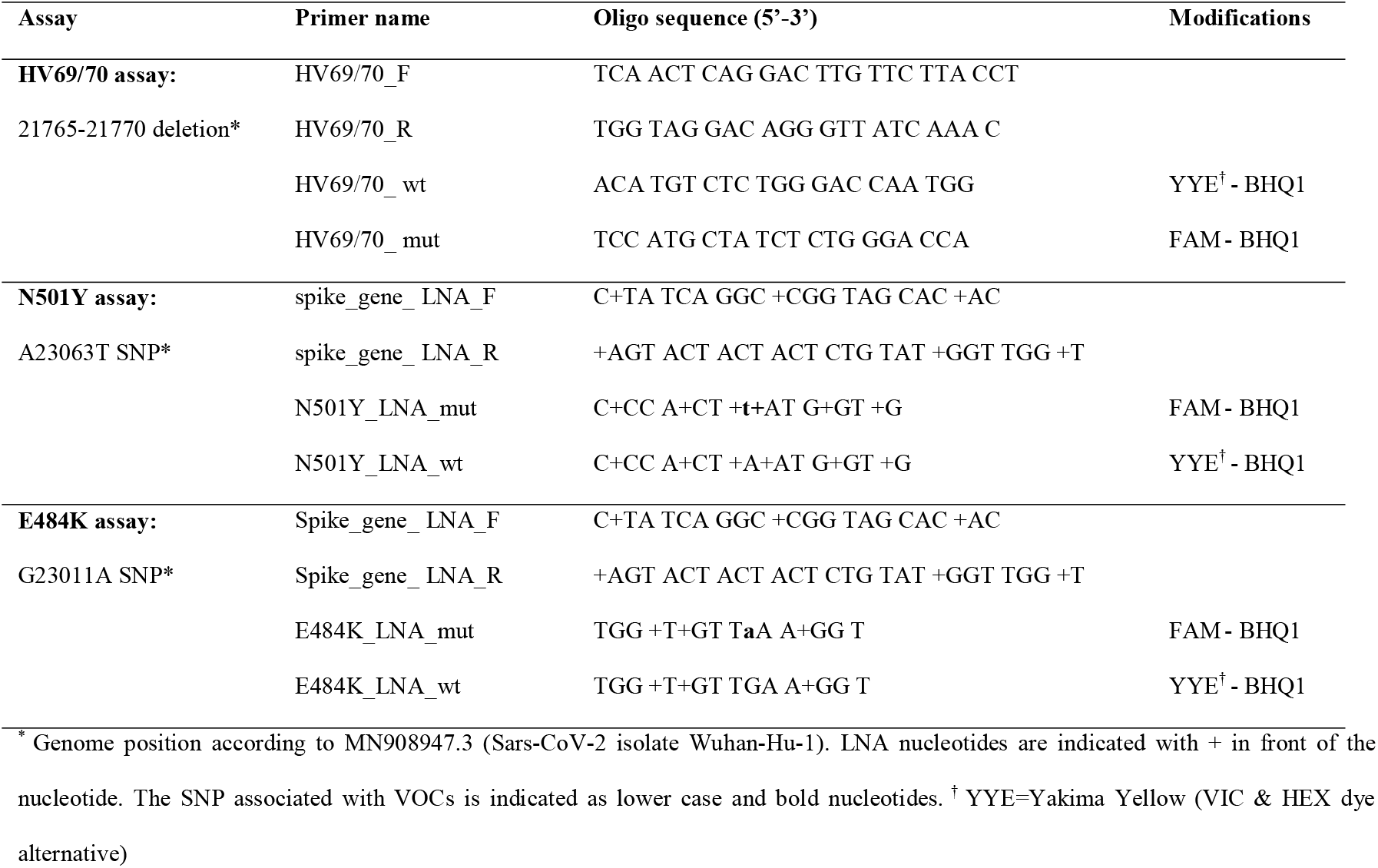
Primer and probe combinations developed for Sars-CoV-2 VOC identification and discrimination.

### Sars-CoV-2 HV69/70-, E484K-and N501Y-specific RT-qPCR assays

The HV69/70-, E484K-and N501Y-specific assays were performed using the Bio-Rad CFX96 Real-Time PCR System (Bio-Rad Laboratories, California, USA). A RT-qPCR run was completed within 1 hour and 10 min using the following thermal profile: reverse 7 transcription step at 50 °C for 5 min; polymerase activation 95 °C for 20 s; and 45 cycles of 3 s at 95 °C and 30 s at 61 °C. Each reaction consisted of 2 μL RNA and 8 μL reaction master mix containing 1x TaqMan™ Fast Virus 1-Step Master Mix (Thermo Fisher Scientific, Leiden, The Netherlands) and the corresponding 1x primer/probe mixture consisting of 0.4 µM primers and 0.2 µM probes. All RT-qPCR assays were run in duplicates with appropriate controls. The mutated sequences were detected by FAM-labelled probes and the wildtype sequences by Yakima-Yellow-labelled probes in multiplex reactions. Data analysis of the RT-qPCR data was conducted using CFX Maestro Software (Bio-Rad Laboratories, California, USA).

The HV69/70-, E484K-and N501Y-specific RT-qPCR assays were transferred to the *peak*PCR platform (Diaxxo AG, Zurich, Switzerland) on which FAM-labelled probes detected the mutated sequence variations only. In order to simplify the testing procedure for the user, the *peak*PCR aluminium sample holders (herein referred to as chips) were preloaded with all necessary reagents in a freeze-dried form. Lyophilized chips were loaded with 4.4 µL sample, sealed off with 1.2 mL of paraffin oil (Sigma–Aldrich, Germany) and run on the *peak*PCR device using the following program: reverse transcription step at 50 °C for 5 min; initial denaturation at 95 °C for 60 s; and 45 cycles of 6 s at 95 °C and 30 s at 62 °C. Total runtime of a *peak*PCR experiment was 37 minutes. *Peak*PCR data was analysed using the *peak*PCR *data*Analysis 1.0 software (Diaxxo AG, Zurich, Switzerland). No drop in performance was observed when lyophilized reagents were used compared to non-lyophilized standard RT-qPCR reagents (Appendix figure 2).

## Results

### Design of RT-qPCR assays for rapid identification of Sars-CoV-2 VOCs

Three mutation-specific RT-qPCR assays based on TaqMan^®^ chemistry were designed. The first assay targets the 6 bp deletion in the spike gene leading to the loss of two amino acids at positions 69 and 70 within the spike protein (HV69/70 assay). This deletion is found in the B.1.1.7 lineage, but not in B.1.351 and P.1. Universal primers amplify a 102-bp (wildtype) or 96-bp (mutant) amplicon. Based on the presence or absence of the deletion, either a FAM-labelled probe or YYE-labelled probe binds and the resulting fluorescence is detected. The second assay targets a non-synonymous SNP in the spike gene (A23011G) leading to an amino acid exchange at positions 484 (E484K). The E484K mutation is only present in VOCs lineages P.1 and B.1.351. In a multiplex reaction, a YYE-labelled probe detects the wildtype and a FAM-labelled probe the mutated sequence. The third assay targets a non-synonymous SNP in the spike gene (A23063T) leading to an amino acid exchange at positions 501 (N501Y). The N501Y mutation is present in all three VOCs but not in non-VOCs. Similar to the E484K assay, a YYE-labelled probe detects the wildtype and a FAM-labelled probe the mutated sequence. A summary of the oligonucleotide sequences used are provided in Table 1.

The novel assays were run on two different RT-qPCR platforms in parallel. On the Bio-Rad CFX96 platform (Figure 1A), the three sequence-discriminatory assays were run as duplex assays, detecting the wildtype sequence in the YYE channel (Figure 1B) and the mutated sequence in the FAM channel (Figure 1C). As a second technology platform the Diaxxo *peak*PCR device was selected (Figure 1D), which is a portable and rapid diagnostic technology platform running the RT-qPCR reaction on ready-to-use chips (Figure 1E). Fluorescence is detected using the Raspberry Pi Camera Module V2 as an inexpensive CCD sensor (Figure 1F). For the *peak*PCR device the multiplex assays were reduced to mutation-specific assays, capable of detecting the mutated sequences only.

**Figure 1.**
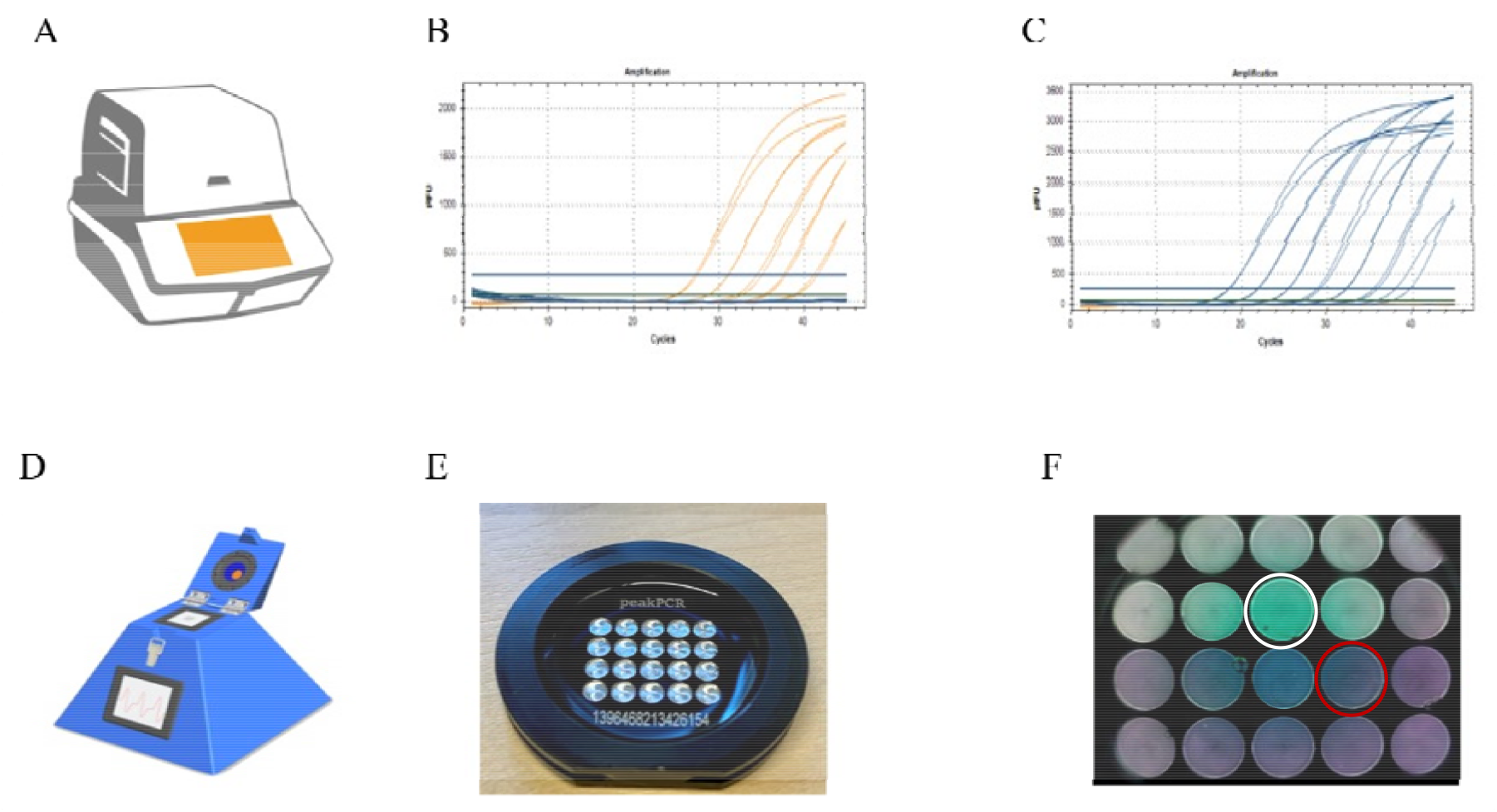
Two RT-qPCR platforms for Sars-CoV-2 VOC identification. A) Standard RT-qPCR device Bio-Rad CFX96. B) Detection of N501Y-wt in serial dilution of Wuhan Hu-1 lineage, ranging from 1-10’000 cp/µL using the YYE-channel of the Bio-Rad CFX96 instrument. C) Detection of N501Y-mut in serial dilution of P.1 lineage, ranging from 1-1’000’000 cp/µL using the FAM-channel of the Bio-Rad CFX96 instrument. D) Portable and rapid diagnostic platform peakPCR. E) Ready-to-use chips with preloaded lyophilized RT-qPCR reagents. F) Photo after cycle 45 detecting fluorescence in each well with an CCD sensor. The depicted well marked with a white circle contains a positive signal after RT-qPCR amplification, while the negative sample, marked with a red circle, did not display an amplification of a PCR product.

### Analytical performance of HV69/70, E484K, and N501Y assays using well-characterized RNA from Sars-CoV-2 VOCs

Four Sars-CoV-2 lineages, namely Wuhan-Hu-1, B.1.1.7, B.1.351 and P.1, were used to assess the RT-qPCR efficiency, specificity and sensitivity of the novel mutation-specific assays. We used serial dilutions, ranging from 1 to 1,000,000 cp/µL, of cell culture derived viral RNA for assay characterization (raw data is provided in *Appendix figure 3*). The integrity of RNA molecules in these serial dilutions was confirmed by monitoring the pan-Sarbecovirus E-gene amplification. The RT-qPCR efficiency was above 80% for all assays on both platforms which translates into moderate to high RT-qPCR efficiencies (Figure 2A).

**Figure 2.**
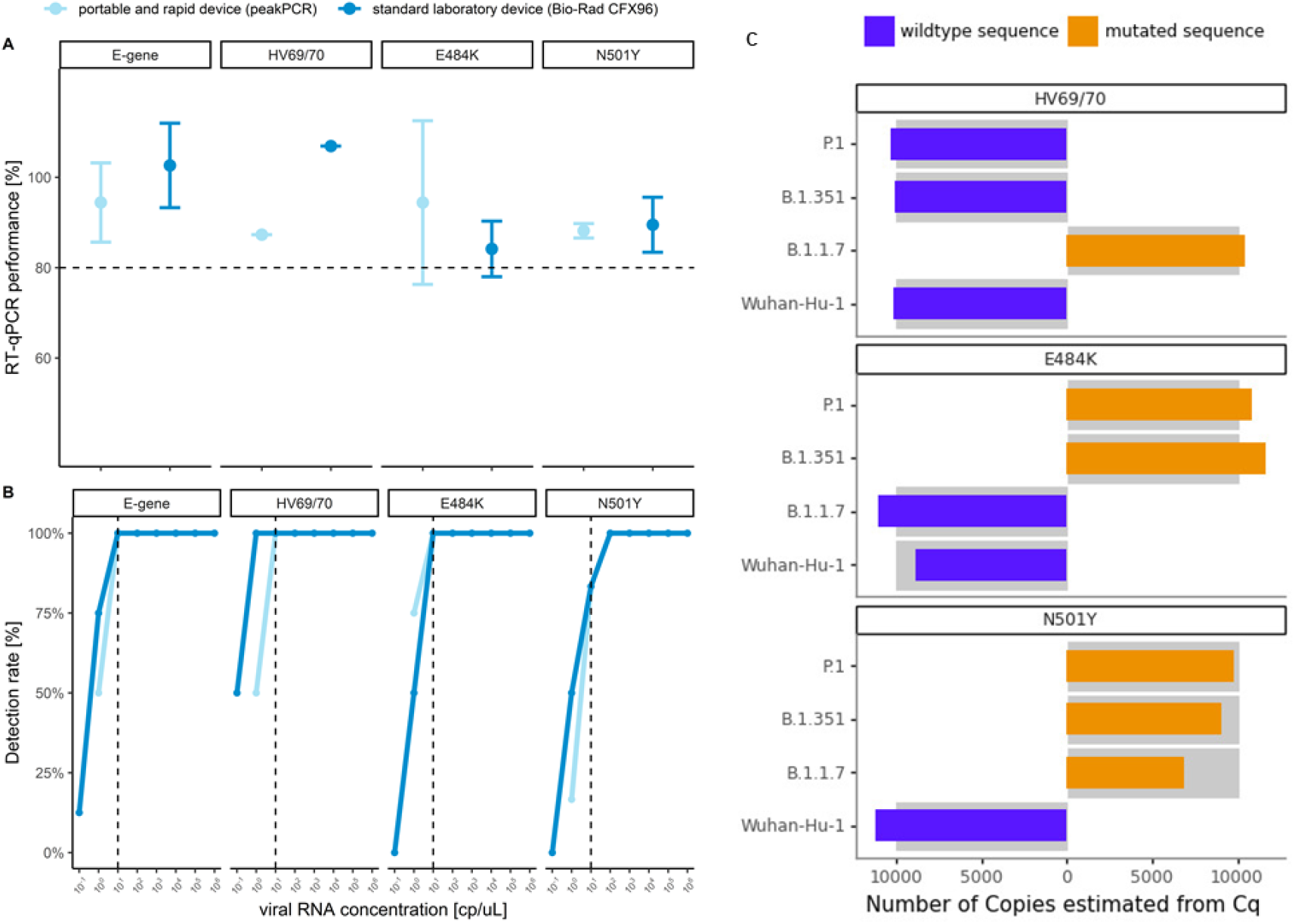
Analytical performance of HV69/70, E484K, and N501Y detecting RT-qPCR assays. A) RT-qPCR efficiency of the E-gene, HV69/70, E484K and N501Y assays as determined by serial dilutions of RNA derived from four cell culture supernatant Sars-CoV-2 lineages. B) Analytical sensitivity represented by detection rates calculated from all replicates for a each viral RNA concentration. The dashed line represents the limit of detection of 10 copies per µL. C) Analytical specificity for the multiplex sequence-discrimination assays run on the Bio-Rad CFX96 device. The data shown are based on RT-qPCR amplification for viral RNA concentrations of 10’000 cp/µL. The grey bar represents the theoretical concentration of 10’000 cp/µL.

The analytical sensitivity of the assays was defined as the lowest viral RNA concentration at which mutations are identified with high confidence. We used the detection rate among all replicates combined for all four Sars-CoV-2 lineages to identify the limit of detection (LOD) (Figure 2B). For the E-gene, HV69/70, and E484K assay a detection rate of 100% was achieved at viral RNA concentration as low as 10 cp/µL. At the same concentration for the N501Y assay, 5 out of 6 replicates (83%) were amplified. At the LOD of 10 cp/µL (dashed line in Figure 2B) no difference between the two RT-qPCR platforms in terms of sensitivity was observed. Viral RNA concentrations below 10 cp/µL cannot be detected, with exception of the HV69/70 assay run on the Bio-Rad CFX96 device, where 1 cp/µL is still reliably detected.

Specificity of all three assays and their ability to distinguish between mutated and wildtype sequences were assessed by testing the assays with RNA from Sars-CoV-2 cell culture supernatants. On both platforms, no signal was observed at any viral RNA concentration if there was not a perfect sequence match of the oligos to the sequence to be detected, resulting in a 100% analytical specificity (*Appendix figure 2*). At a viral RNA concentration of 10’000 cp/µL, the HV69/70, E484K, and N501Y genotypes were by the wildtype-mutant multiplex assays run on the Bio-Rad CFX96 platform correctly identified (Figure 2C). The mutation-specific probe of the HV69/70 assay gave a signal only when run with RNA of the B.1.1.7 lineage carrying the mutation. The E484K-mutation assay did not result in amplification when run on RNA from Wuhan Hu-1 and B.1.1.7 lineage. The N501Y-mutation assay detected correctly all VOCs but not the Wuhan Hu-1 lineage. In summary, the three assays correctly identify lineage-associated mutations with moderate to high RT-qPCR efficiencies in samples with more than 10 cp/µL of Sars-CoV-2 RNA. We also demonstrated that these assays can be successfully conducted on the rapid diagnostic platform *peak*PCR and the performance in terms of sensitivity and specificity does not significantly differ between these two RT-qPCR platforms.

## Discussion

The COVID-19 pandemic is a continuous, unprecedented, global public health crisis with severe economic and social consequences. More than one year into the pandemic, the emergence of VOCs starts to pose again serious threat to contain the virus. Rapid and reliable identification of SARS-CoV-2 variants is a critical component of public health interventions to mitigate the further spread of VOCs that might undermine performance of diagnostic tests and vaccine induced immunity against this virus.

We designed, tested and validated three mutation-specific RT-qPCR assays, detecting the E484K and N501Y SNPs as well as the 6-bp deletion affecting HV69/70, all located in the Sars-CoV-2 spike gene. All assays can be performed under standard RT-qPCR conditions simplifying integration into existing laboratory environments and proved to be highly sensitive, specific, and reproducible.

The approach presented here is well-suited for cost-effective, robust and high-throughput screening of large cohorts. Mutation-specific RT-qPCRs are not intended to replace NGS, but rather complement and extend molecular surveillance programs and focal outbreak monitoring. As demonstrated, the perfect mismatch discrimination of LNA-based assays results in 100% specificity and the simplicity of this type of assay in both, design and implementation, will allow rapid adaptation (within few days) to new VOCs which will be identified in future. A similar LNA-based approach for detection of N501Y and HV69/70 has been successfully tested in Canada on 2,430 samples ^18^. The results of their in-house assay were concordant with the commercial assay VirSNiP SARS-CoV-2 (TIB Molbiol, Berlin, Germany) which is based on melting-curve analysis. This underlines the possibility of using LNA-based RT-qPCR assays for single-nucleotide discrimination as opposed to using melting-curve analysis. Although, Sars-CoV-2 SNP genotyping by melting-curve analysis is widely used ^19^, the LNA-based approach has several advantages: It is faster, easier to integrate into existing laboratory workflows and could be combined with an diagnostic assay and therefore allow immediate genotyping. Furthermore, LNA-based sequence-discriminatory assays are better suited to identify coinfections of more than one lineage of SARS-CoV-2, a phenomenon which was recently observed in Brazil ^20^.

In order to reduce the sample-to-result turnaround time our strategy included the transformation of the assays to a portable, robust and rapid diagnostic RT-qPCR platform and therefore allow decentralization of testing. The *peak*PCR platform completed the sample analysis in 37 minutes, which is half of the time required to run the same assay on a standard RT-qPCR platform while retaining a similar efficiency, specificity and sensitivity. Apart from the speed, also the simplicity of usage and, relatively low costs and the independence from cold chains for reagent supply are significant advantages which make this platform well-suited for decentralized, rapid testing particularly in resource limited settings.

## Data Availability

The authors confirm that the data supporting the findings of this study are available within the article and its supplementary materials.

## Contributors

Conceptualization by SH, PB, PW and TS. *peak*PCR platform technology and chip design: PB, ARS, MG, WJS. Formal analysis was done by PW and TS. DS and OE provided the cell culture derived RNA from Sars-CoV-2 VOCs. The first draft of manuscript was written by PB, CAD and TS. All authors reviewed and approved the final manuscript.

## Disclaimers

PB, MG and WJS are co-inventors on a corresponding patent application around the detection platform and shareholders of the ETH spin-off company Diaxxo AG. TS has consulted for Diaxxo AG. All other authors declare no competing interests.

## Appendix

**Appendix data 1.**
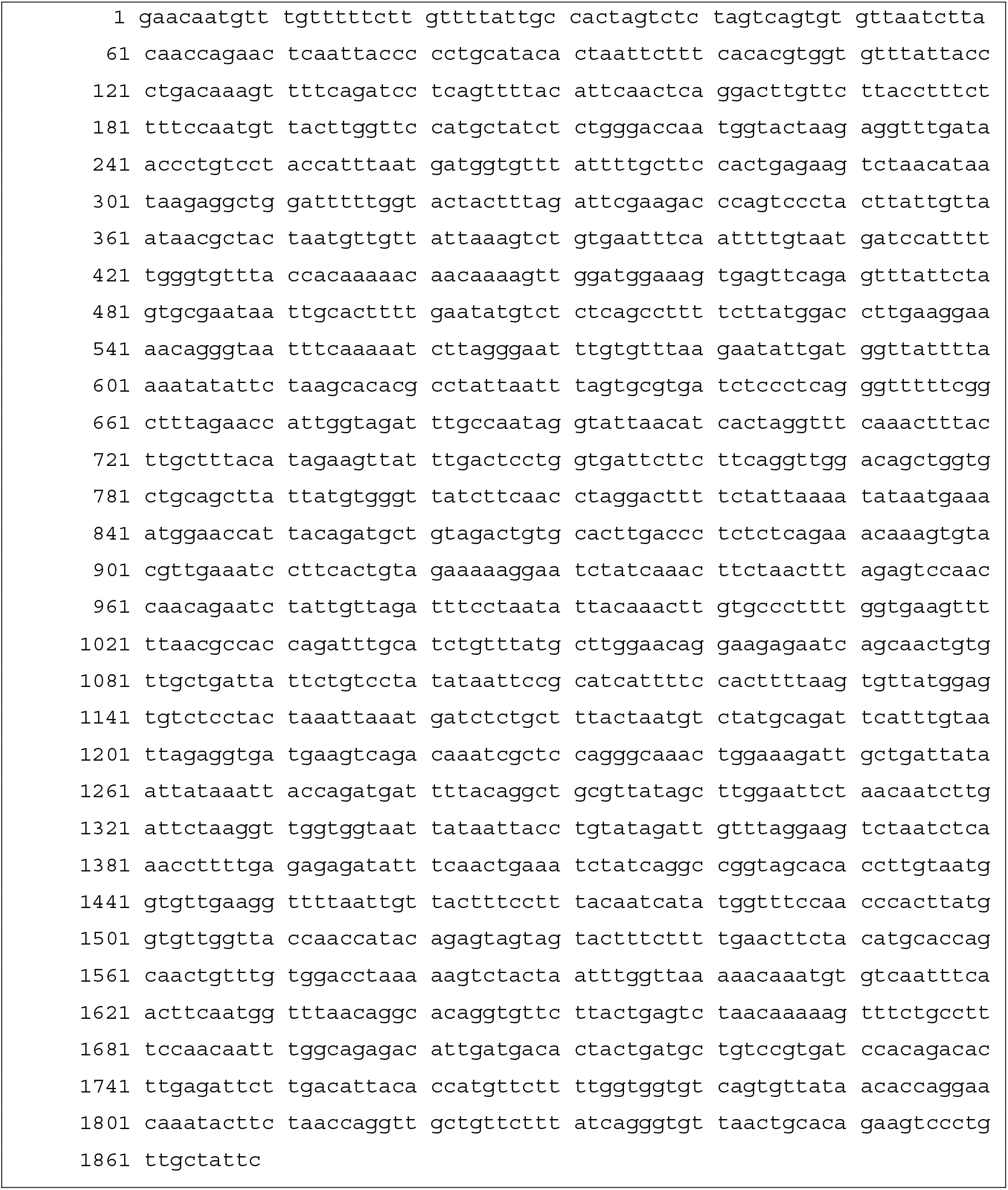
A 1869 bp long synthetic Sars-CoV-2 spike gene fragment (genome position 21,557-23,434 bp), based on the sequence of B.1.1.7 was synthesized by Eurofins Genomics, Konstanz, Germany.

**Appendix figure 1.**
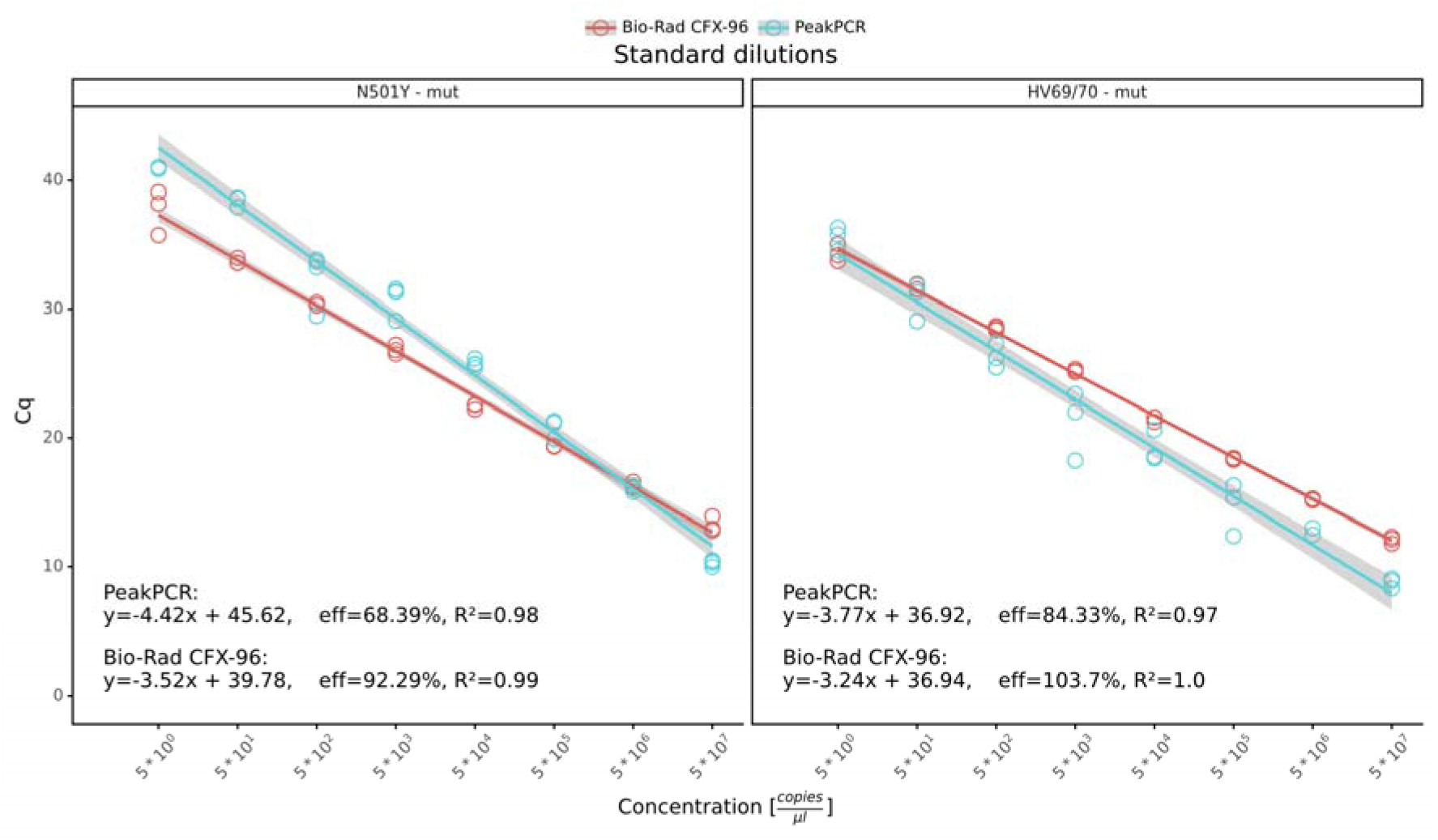
Synthetic spike gene serial dilution experiments for HV69/70 and N501Y assays.

**Appendix figure 2.**
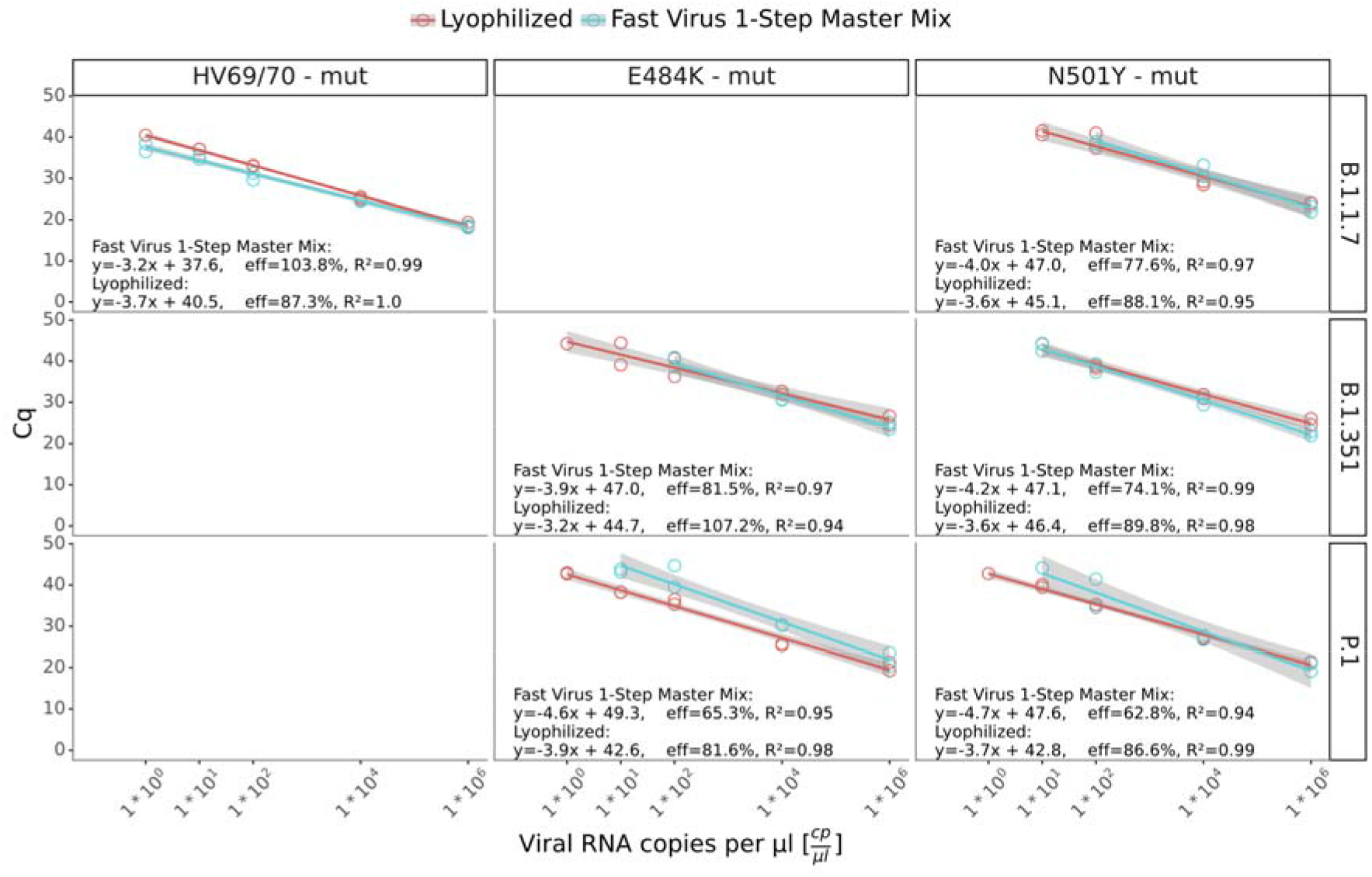
Comparison of lyophilized reagents with non-lyophilized standard RT-qPCR reagents on the *peak*PCR device

**Appendix figure 3.**
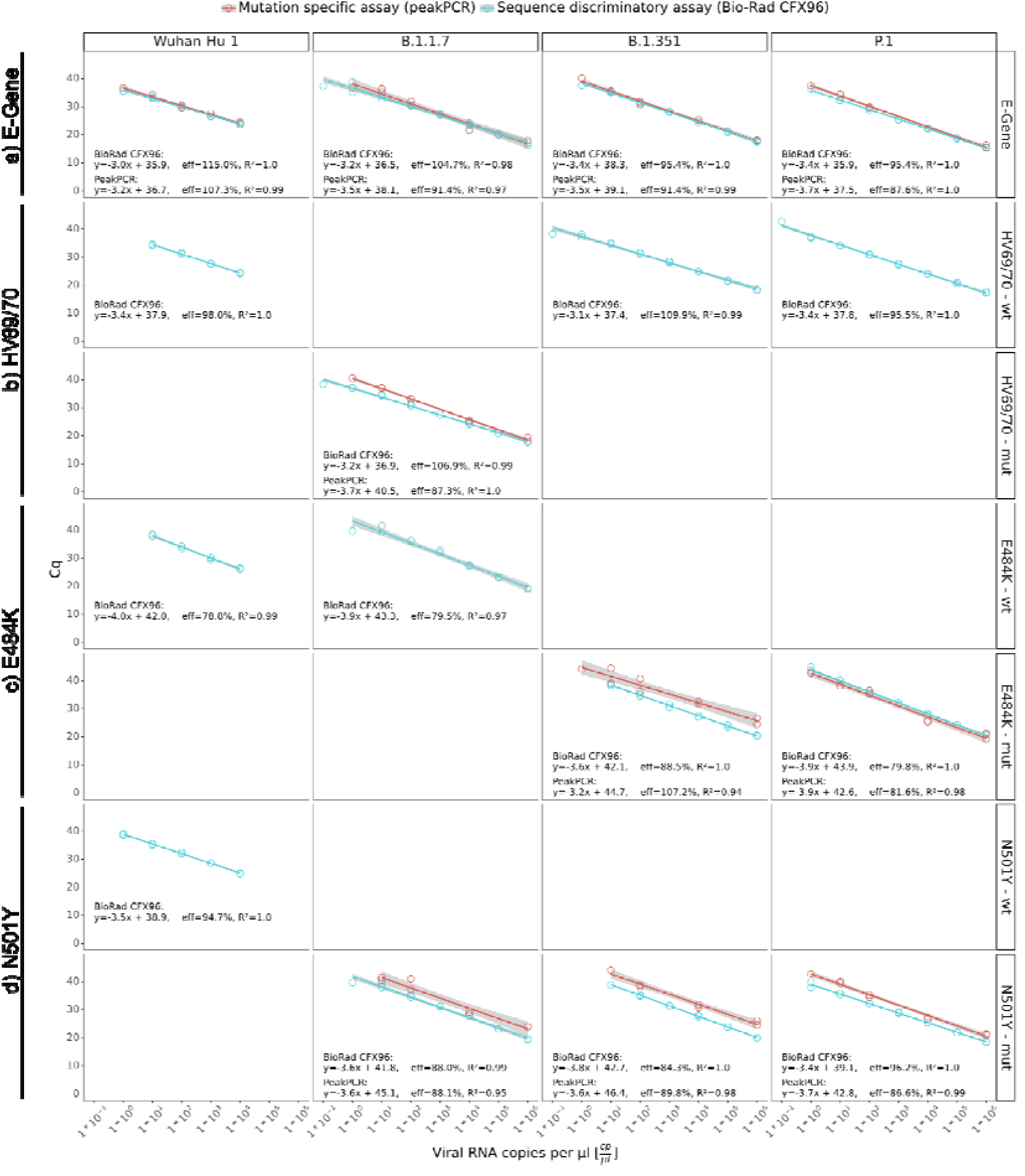
RT-qPCR performance of novel Sars-CoV-2 mutation-specific assays. a) Sars-CoV-2 E-gene reference assay b) HV69/70 assay, c) E484K assay and d) N501Y assay. Each circle represents a technical replicate. With exception of the HV69/709-wt assay, wild-type assays were only run on the Bio-Rad CFX96 platform. For the Wuhan-Hu-1 lineage, the two highest RNA concentrations of 1,000,000 and 100,000 cp/µL were not available. Performance experiments on the peakPCR device used the 1,000,000 cp/µL, 10,000 cp/µL, 100 cp/µL, 10 cp/µL, and the 1 cp/µL concentrations.

